# ASSESSMENT OF PATIENT SAFETY ATTITUDE AMONG DOCTORS AND NURSES AT PUBLIC SPECIALIST HOSPITALS IN MALAYSIA

**DOI:** 10.1101/2024.08.17.24311962

**Authors:** Sharifah Balqis Sayed Abdul Hamid, Aniza Ismail, Saperi Sulong

## Abstract

Patient safety remains a global problem that affects both developed and developing countries. Healthcare organizations should focus on the need of assessing safety culture as that will provide basic understanding on safety related perceptions of their staffs.

**Objective:** This study was aimed to assess baseline level and mean score of every domain of the patient safety attitude among doctors and nurses in public hospitals in Selangor and to identify the determinants associated with patient safety attitude in all domains : teamwork, safety climate, working condition, job satisfaction, stress recognition and perception of management.

**Method:** This was a cross-sectional study utilizing the Safety Attitude Questionnaires (SAQ) involving 142 doctors and 231 nurses in three public hospitals in Selangor. The samples were selected through proportionate stratified random sampling. Data was analysed using descriptive, bivariate and multivariate analysis.

**Results:** The response rate was 83% in which job satisfaction (73.78±20.54) and perception of management (58.98±16.28) received the highest and lowest mean score, respectively. The factors associated with positive patient safety attitude towards 1)teamwork were position (OR 1.99, p=0.03) and history of attending patient safety training (OR 3.228, p = 0.000); 2) safety climate were history of attending patient safety training (OR 7.283, p = 0.000); 3) job satisfaction were education level (OR 35.709, p=0.000), position (OR 10.598, p=0.000) and history of attending patient safety training (OR 2.883, p = 0.000); 4) perception of management were age (OR 2.084,p=0.021) and work area (OR 2.461,p=0.012); 5) working condition were age (OR2.200,p 0.003) and history of attending patient safety training (OR1.738, p 0.032).

**Conclusion:** Respondents with history of attending patient safety training have more positive attitude towards teamwork, safety climate, job satisfaction and working condition. Improving patient safety culture should be priorotized by the hospital management team to inculcate safety culture in healthcare providers.

**STRENGTHS AND LIMITATIONS OF THIS STUDY:**

- This study reports the outcomes of patient safety attitude in public hospitals in Malaysia.
- This study has a good response rate.
- The study covers both types of hospitals in Malaysia (specialist and non-specialist hospitals).
- The respondents were from 2 categories of healthcare professionals.
- The factor of limited time and financial constraint limit the ability to include bigger sample into this study

## BACKGROUND

The Hippocratic Oath, a foundation of medical practice, urges practitioners to “first, do no harm.” However, the 1999 Institute of Medicine (IOM) report, To Err Is Human: Building a Safer Health System revealed that much harm was being done. This now often cited report compiled statistics suggesting that as many as 98,000 people may be dying each year as a result of medical errors. Medical facilities have long had systems in place to monitor errors. These systems, however, mostly consisted of the filing of incident reports once an error was discovered. [1,2]

Retrospective approach that often-assigned blame to individuals and did little to analyze systems, identify trends, or make recommendations for overall improvements. With increasing media attention and public awareness of medical errors, it became clear that this system of monitoring was inadequate. Patient safety has become a central focus for most medical institutions and many new programs to monitor safety and prevent medical mistakes have emerged as a result. The use of new technologies, the employment of automated systems, the introduction of system redundancies, the use of event simulations, and the implementation of new staff training are all strategies that have been put in place in an attempt to reduce the rate of errors. [1]

Malaysian Patient Safety Goals (MPSG) were developed by the Patient Safety Council of Malaysia with Patient Safety Unit, Ministry of Health as the Technical Secretariat and relevant stakeholders to outline key priority areas on patient safety and challenge our healthcare organizations to improve some of the most significant and challenging patient safety issues in Malaysia. These goals which have been implemented since 2013, are applicable to both public and private health care facilities in Malaysia. The Malaysian Patient Safety Goals will allow systematic monitoring and evaluation of patient safety status in Malaysia. The first version of these goals included 15 goals and 59 KPIs. Subsequently, these were then reduced to 13 safety goals and 19 KPIs which are more manageable. The goals are based on WHO key priority areas, Joint Commission International, Patient Safety Goals and key patient safety issues in Malaysia. [3]

All healthcare facilities in Malaysia need to monitor their performance regularly and take actions for improvement. Only the overall annual performance needs to be submitted to Patient Safety Unit, Ministry of Health via online system e-goals patient safety. MPSG has been implemented since July 2013 in all healthcare facilities nationwide. Many efforts have been made by Patient Safety Unit, Ministry of Health Malaysia to ensure its implementation in public hospitals. These include formulation of policy document from Director General of Health Malaysia No 2/2013, formulation and launching of the MPSG Guideline, making MPSG as part of MSQH hospital accreditation standards, establishment of user-friendly online e-goals patient safety reporting system, extensive training from top leaders to staff, presentations in conferences/seminars, formulation of reports which is easily assessable online, establishment of Mandatory Patient Safety Course for House Officers which include MPSG as part of the subjects, formulation of Specific Technical Working Group for each goals, extensive discussions and engagement sessions with staffs, promotional activities and also delivering of MPSG Report Card by Director General of Health Malaysia to each state health departments. [3]

The Malaysian Patient Safety Goals 2003 was implemented to address patient safety issues in public and private health facilities in the nation. For instance, one of the goals identified was the need to develop a medical error reporting system that promoted a safe environment by adopting a ‘reporting and learning’ as well as ‘just and non-blaming’ culture. For example, in an outpatient geriatric pharmacy in Malaysia, 20 cases of medication errors were reported to occur daily, and the estimated cost of the medication errors was Malaysian Ringgit MYR301 (£54) daily or MYR9327 (£1667) a month and approximately MYR111 924 (approximately £20 000) a year [4] Challenges in implementation of patient safety goals are many, as safety encompasses cultural, behavior, technical, clinical and psychological domains. In order to transform the cultural aspect of safety, there is a need to acknowledge and to understand it. Measuring the patient safety attitude is essential to determine predictor factors that influence patient safety attitude. One way to aid healthcare leaders in understanding their organizations safety culture is to administer a survey using safety attitude assessment tools. [5,6] These tools can be utilized to evaluate the relationships between safety attitude domains and the associated factors.

Statistics have shown that all MOH hospitals implement MPSG. Nevertheless, it is not known how far the implementation has managed to give an impact to our healthcare facilities in terms of improving our Patient Safety Culture. Hence, it is essential for this research to be conducted to assess the Safety Attitude Perception among doctors and nurses in a few identified hospitals. Good Patient Safety Culture in our healthcare facilities are indicated by better Patient Safety Attitude among the healthcare providers. Because healthcare providers are critically responsible for optimization and prevention of any harmful events or possibility to cause harm to patients, a study on patient safety attitude of healthcare providers will provide an insight towards their attitude and assist in identifying specific areas for improvement. Consequently, a safe culture that is targeted to reduce errors can be further engineered into daily work practices. Thus, this study’s main objective was to assess baseline level and mean score of each patient safety attitude domain among doctors and nurses in public hospitals in Selangor and identify the determinants associated with patient safety attitude in all domains.

## METHODS

### Study Design and Sampling

This cross-sectional study was conducted at three public hospitals, whereby they implement similar patient safety practices and policies. Data were collected from July 2019 to January 2020. All doctors and nurses who were involved directly with patient care processes and who had been working at the hospitals for at least 4weeks were included in the study. Those who worked in management and who were on a long leave were excluded from the study.

The samples were selected through proportionate stratified random sampling to ensure that, throughout the population, the sample size selected from each subgroup was proportional to the size of that subgroup. The same sampling method was used to determine how many representatives from each professional category would be selected. The sample size required, which was calculated manually using Kish L. Formula, was 382 at 95% CI and with 80% power. However, considering a dropout rate of 20%, the final sample size required was 458.

### Measures

This study has been done using validated questionnaire. One of the most rigorously validated and commonly used tool for measuring safety culture in healthcare is the Safety Attitudes Questionnaire (SAQ). The tool has been adapted for use in intensive care units (ICUs), general inpatient settings such as medical and surgical wards, emergency medical services, operation theatres, ambulatory clinics or primary care, community pharmacies and nursing homes [5]

This questionnaire has well constructed validity and internal consistency, as well as good psychometric properties, and is associated with clinical and patient outcomes.[4,5] The SAQ is a 60-item questionnaire with closed-ended responses asking the respondents to indicate their level of agreement with each item on a 5-point scale ranging from ‘1’ (strongly disagree) to ‘5’ (strongly agree). There are several versions developed for different healthcare settings. All versions consist of 30 identical core questions, eliciting respondents’ attitudes through six domains: Teamwork Climate, Safety Climate, Perceptions on Management, Job Satisfaction, Working Conditions and Stress Recognition, using a 5-point Likert scale. For example, six individual items, when taken together, comprised a respondent’s perception of Teamwork Climate.

In this study, the survey was based on the 36 core questions of the SAQ (short form), rather than the 60-item questionnaire. The reasons being that validation and benchmarking data have only been published for the core items. [5] Additionally, the SAQ was adapted and translated to the Malay version (MSAQ) from the generic version (Short Form 2006), which contains the six safety dimensions mentioned above. The process of adaptation included forward translation and backward translation method, followed by content validity analysis by seven subject matter experts. All 36 items of the SAQ were retained for the field test. The Malaysian SAQ (MSAQ) was distributed to 400 healthcare workers in a hospital in Kuala Lumpur. [7]

In addition, the information on the number of self-reported error (including near misses) that respondents have done in the past 12months in their current working institution was also obtained. On the instruction section of the front page, the following definition of medication error will be given: ‘An error is any type of medication error, mistake, incident, or quality related event, regardless of whether or not (near miss) it reaches the patient or results in patient harm. Errors may be related to, or include: prescribing, transcribing, dispensing, administering, monitoring (use of medication), supplying, giving information, preparing, unsafe conditions or procedures in the pharmacy’. Respondent’s demographic information (age, gender, race, education level, position and income). Meanwhile, number of years of work experience, field of working area and number of safety training attended also obtained.

Data were analysed using SPSS V.21, and the respondents’ demographic characteristics and patient safety attitude level were determined using univariate analysis. Before the analysis, three negatively worded items (items 2, 11 and 36) in the SAQ were reversed. Each item’s score was calculated by converting the 5-point Likert scale into a 100-point scale: 1=0, 2=25, 3=50, 4=75 and 5=100. Each item’s score within the same dimension was summed and divided by the number of items available for that dimension to obtain a score of 0–100. If a respondent’s mean score was ≥75, they had a positive safety culture for a given dimension. The respondent’s overall score for the patient safety attitude level was calculated using the sample method. Descriptive statistic has been performed to describe the sociodemographic characteristics. Simple and multiple logistic regression has been performed to calculate Odd ratio and also adjusting for the confounding factors. This is to estimate the association of the factor towards patient safety attitude. In this study, consider p value less than 0.05 to be statistically significant, with 95% of confident interval.

## RESULTS

A total of 450 questionnaires were distributed to doctors and nurses in three selected hospitals during several visits. Three hundreds and seventy three respondents completed and returned the questionnaires, resulting in an overall response rate of 83%.

## DESCRIPTIVE ANALYSIS

### Demographic Characteristic

Table 1, represents the background of the respondents participated in the study. Most respondents were female and Malay aged less than 30 years old. Majority of them were nurses (62%), had lower income, diploma holder with working experience of 1-5 years. Almost half of the respondents were working in medical area, most of them (63.5%) have attended patient safety training and As for the numbers of self-reported events in twelve months, 64% of the respondents did not report any events followed by 33% of the respondents reported 1-10 events while only 3.0% of the respondents reported more than ten events.

**Table 1.** Demographic characteristics of respondents.

| Characteristics | Respondents (n= 373) |  |
| --- | --- | --- |
|  | N | Frequency (%) |
| Age (years) |  |  |
| Age Group |  |  |
| Below 31 | 209 | 56 |
| 31-40 | 134 | 36 |
| 41-50 | 28 | 7.5 |
| More than 50 | 2 | 0.5 |
| Gender |  |  |
| Male | 54 | 14.5 |
| Female | 319 | 85.5 |
| to be continued... |  |  |
| ...continuation |  |  |
| Race |  |  |
| Malay | 289 | 77.5 |
| Chinese | 25 | 7 |
| Indian | 43 | 11.5 |
| Others | 16 | 4 |
| Education level |  |  |
| Diploma | 214 | 57 |
| Degree | 134 | 36 |
| Master/PHD | 25 | 7 |
| Position |  |  |
| Doctor | 142 | 38 |
| Nurse | 231 | 62 |
| Income |  |  |
| Lower | 189 | 50 |
| Middle | 163 | 44 |
| High | 21 | 6 |
| Length of Working |  |  |
| Less than 1 year | 23 | 6 |
| 1-5 years | 181 | 48.5 |
| 6-10 years | 90 | 24 |
| 11-15 years | 43 | 11.5 |
| More than 15 years | 36 | 10 |
| Field of Working Area |  |  |
| Medical | 162 | 43 |
| Surgical | 136 | 37 |
| Others | 75 | 20 |
| History of Attending Safety Training |  |  |
| Yes | 237 | 63.5 |
| No | 136 | 36.5 |
| Number of Self- Reported Error |  |  |
| 0 (Zero Reporting) | 238 | 64 |
| 1-10 | 122 | 33 |
| More than 10 | 13 | 3 |

### Patient Safety Attitude Score

Table 2 demonstrates patient safety attitude among doctors and nurses towards each domain with the respective scoring. The job satisfaction domain received the highest mean score among doctors and nurses (73.78±20.54). In contrast, perception of management was perceived as the least domain, with the lowest mean score (58.98±16.28). In decreasing order, the percentage of doctors and nurses who held positive attitude towards each domain was 62.7% (job satisfaction), 50.1% (stress recognition), 45% (teamwork), 38.1% (safety climate), 35.1% (working condition) and 23.6% (perception of management).

**Table 2.** Patient Safety Attitude.

| Table 2 Patient Safety Attitude |  |  |  |  |
| --- | --- | --- | --- | --- |
| Domain | Minimum | Maximum | Mean (SD) | Positive Response ($\geq 75$) (%) |
| Teamwork | 16.67 | 100.00 | 70.70 (14.85) | 45.0 |
| Safety Climate | 17.86 | 100.00 | 69.57 (13.53) | 38.1 |
| Job Satisfaction | 0.00 | 100.00 | 73.78 (20.54) | 62.7 |
| Stress Recognition | 0.00 | 100.00 | 67.49 (23.73) | 50.1 |
| Perception of Management | 10.00 | 100.00 | 58.98 (16.28) | 23.6 |
| Working Condition | 6.25 | 100.00 | 60.74 (0.85) | 35.1 |

\*Percent positive scores are computed as the percent of respondents who answered 'agree slightly' or 'agree strongly' on each of the items within a scale (ie, 4 or 5 on the original 5-point Likert scale)

**Table 3.** Associated Factors of Patient Safety Attitude on Teamwork.

| Domain | Teamwork |  | X<sup>2</sup> | p value |
| --- | --- | --- | --- | --- |
|  | Positive n(%) | Negative n(%) |  |  |
| Characteristic |  |  |  |  |
| Age (years) |  |  | 0.432 | 0.511 |
| Below 30 | 91 (43.5) | 118 (56.5) |  |  |
| 30 and above | 77 (47.0) | 87 (53) |  |  |
| Gender |  |  | 2.820 | 0.093 |
| Male | 30 (55.6) | 24 (44.4) |  |  |
| Female | 138 (43.3) | 181 (56.7) |  |  |
| Race |  |  | 2.112 | 0.146 |
| Malay | 136 (47.1) | 153 (52.9) |  |  |
| Non- Malay | 32 (38.1) | 52 (61.9) |  |  |
| Education level |  |  | 3.179 | 0.204 |
| Diploma | 88 (41.1) | 126 (58.9) |  |  |
| Degree | 68 (50.7) | 66 (49.3) |  |  |
| Master/PHD | 12 (48) | 13 (52.0) |  |  |
| Position |  |  | **5.602** | **0.018** |
| Doctor | 75 (52.8) | 67 (47.2) |  |  |
| Nurse | 93 (40.3) | 138 (59.7) |  |  |
| Income |  |  | **4.443** | **0.035** |
| < RM 3860 | 75 (39.7) | 114 (60.3) |  |  |
| RM3860 and above | 93 (50.5) | 91 (49.5) |  |  |
| Length of Working |  |  | 0.677 | 0.954 |
| Less than 1 year | 10 (43.5) | 13 (56.5) |  |  |
| 1-5 years | 83 (45.9) | 98 (54.1) |  |  |
| 6-10 years | 41 (45.6) | 49 (54.4) |  |  |
| 11-15 years | 17 (39.5) | 26 (60.5) |  |  |
| More than 15 years | 17 (47.2) | 19 (52.8) |  |  |
| Field of Working Area |  |  | **8.488** | **0.014** |
| Medical | 67 (41.4) | 95 (58.6) |  |  |
| Surgical | 56 (41.2) | 80 (58.8) |  |  |
| Others | 45 (60) | 30 (40) |  |  |
| History of Attending Safety Training |  |  | **19.179** | **0.000** |
| Yes | 127 (53.6) | 110 (46.4) |  |  |
| No | 41 (30.1) | 95 (69.9) |  |  |
| Number of Self- Reported Error |  |  | 0.369 | 0.544 |
| No (Zero Reporting) | 110 (46.2) | 128 (53.8) |  |  |
| Yes | 58 ( 43) | 77 (57) |  |  |

## BIVARIATE ANALYSIS

In bivariate analysis, position in the hospital, income, field of working area and history of attending patient safety training were identified to have significant association with teamwork domain. For safety climate, only field of working area and history of attending patient safety training were significantly associated. For job satisfaction, race, education level, position, income, field of working area and history of attending safety training were identified as significantly associated. Furthermore, education level, position and field of working area were significantly associated with stress recognition domain. While age, education level, position, income, working area and history of attending training were identified as significantly associated with Perception of Management domain. Lastly, for working condition, age, education level, position, income, field of working area and history of attending patient safety training were significantly associated. As we can see, history of attending patient safety training seemed to have significant association with all domains except for stress recognition.

## MULTIVARIATE ANALYSIS

In single logistic regression, the significant factors for teamwork domain were identified as position in the hospital, income, field of working area and history of attending patient safety training. After running for multiple logistic regression, the determinants of having positive safety attitude towards teamwork were position, work area and history of attending patient safety training (Table 4.9) From this analysis it can be interpreted as: 1) doctors have 2 times the odds to have positive safety attitude towards teamwork compared to nurses (95% CI 1.071-3.702, p=0.03), 2) those who work in medical area have 2 times the odds to have negative attitude towards teamwork compared to non-clinical area (95% CI 0.218-0.762, p=0.005) and surgical area have 3 times the odds to negative attitude towards teamwork compared to non-clinical area (95% CI 0.189-0.642, p=0.001), 3) those who attended patient safety training have 3 times the odds to have positive safety attitude towards teamwork compared to those who do not attend patient safety training.

**Table 4.** Associated Factors of Patient Safety Attitude on Safety Climate.

| Domain | Safety Climate |  | X<sup>2</sup> | p value |
| --- | --- | --- | --- | --- |
|  | Positive n(%) | Negative n(%) |  |  |
| Characteristic |  |  |  |  |
| Age (years) |  |  | 0.962 | 0.327 |
| ...continuation |  |  |  | To be continued.. |
| Below 30 | 75 (35.9) | 134 (64.1) |  |  |
| 30 and above | 67 (40.9) | 97 (59.1) |  |  |
| Gender |  |  | 1.812 | 0.178 |
| Male | 25 (46.3) | 29 (53.7) |  |  |
| Female | 117 (36.7) | 202 (63.3) |  |  |
| Race |  |  | 0.578 | 0.447 |
| Malay | 113 (39.1) | 176 (60.9) |  |  |
| Non- Malay | 29 (34.5) | 55 (65.5) |  |  |
| Education level |  |  | 3.520 | 0.172 |
| Diploma | 90 (42.1) | 124 (57.9) |  |  |
| Degree | 43 (32.1) | 91 (67.9) |  |  |
| Master/PHD | 9 (36) | 16 (64) |  |  |
| Position |  |  | 0.451 | 0.502 |
| Doctor | 51 (35.9) | 91 (64.1) |  |  |
| Nurse | 91 (39.4) | 140 (60.6) |  |  |
| Income |  |  | 0.746 | 0.388 |
| < RM 3860 | 76 (40.2) | 113 (59.8) |  |  |
| RM3860 and above | 66 (35.9) | 118 (64.1) |  |  |
| Length of Working |  |  | 2.471 | 0.65 |
| Less than 1 year | 9 (39.1) | 14 (60.9) |  |  |
| 1-5 years | 66 (36.5) | 115 (63.5) |  |  |
| 6-10 years | 33 (36.7) | 57 (63.3) |  |  |
| 11-15 years | 16 (37.2) | 27 (62.8) |  |  |
| More than 15 years | 18 (50) | 18 (50) |  |  |
| Field of Working Area |  |  | **13.704** | **0.001** |
| Medical | 48 (29.6) | 114 (70.4) |  |  |
| Surgical | 53 (39) | 83 (61) |  |  |
| Others | 41 (54.7) | 34 (45.3) |  |  |
| History of Attending Safety Training |  |  | **55.992** | **0.000** |
| Yes | 124 (52.3) | 113 (47.7) |  |  |
| No | 18 (13.2) | 118 (86.8) |  |  |
| Number of Self- Reported Error |  |  | 0.282 | 0.595 |
| No (Zero Reporting) | 93 (39.1) | 145 (60.9) |  |  |
| Yes | 49 (36.3) | 86 (63.7) |  |  |

**Table 5.** Associated Factors of Patient Safety Attitude on Job Satisfaction.

| Domain | Job Satisfaction |  | X<sup>2</sup> | p value |
| --- | --- | --- | --- | --- |
|  | Positive n(%) | Negative n(%) |  |  |
| Characteristic |  |  |  |  |
| Age (years) |  |  | 0.788 | 0.375 |
| Below 30 | 127 (60.8) | 82 (39.2) |  |  |
| 30 and above | 107 (65.2) | 57 (34.8) |  |  |
| Gender |  |  | 0.418 | 0.518 |
| Male | 36 (66.7) | 18 (33.3) |  |  |
| Female | 198 (62.1) | 121 (37.9) |  |  |
| Race |  |  | **4.971** | **0.026** |
| Malay | 190 (65.7) | 99 (34.3) |  |  |
| Non Malay | 44 (52.4) | 40 (47.6) |  |  |
| Education level |  |  | **26.669** | **0.000** |
| Diploma | 158 (73.8) | 56 (26.2) |  |  |
| Degree | 63 (47) | 71 (53) |  |  |
| Master/PHD | 13 (52) | 12 (48) |  |  |
| Position |  |  | **12.582** | **0.000** |
| Doctor | 73 (51.4) | 69 (48.6) |  |  |
| Nurse | 161 (69.7) | 70 (30.3) |  |  |
| Income |  |  | **8.277** | **0.04** |
| < RM3860 | 132 (69.8) | 57 (30.2) |  |  |
| RM3860 and above | 102 (55.4) | 82 (44.6) |  |  |
| Length of Working |  |  | 7.941 | 0.094 |
| Less than 1 year | 13 (56.5) | 10 (43.5) |  |  |
| 1-5 years | 107 (59.1) | 74 (40.9) |  |  |
| 6-10 years | 57 (63.3) | 33 (36.7) |  |  |
| 11-15 years | 27 (62.8) | 16 (37.2) |  |  |
| More than 15 years | 30 (83.3) | 6 (16.7) |  |  |
| Field of Working Area |  |  | **26.276** | **0.000** |
| Medical | 78 (48.1) | 84 (51.9) |  |  |
| Surgical | 99 (72.8) | 37 (27.2) |  |  |
| Others | 57 (76) | 18 (24) |  |  |
| History of Attending Safety Training |  |  | **26.917** | **0.000** |
| Yes | 172 (72.6) | 65 (27.4) |  |  |
| No | 62 (45.6) | 74 (54.4) |  |  |
| Number of Self- Reported Error |  |  | 3.751 | 0.053 |
| No (Zero Reporting) | 158 (66.4) | 80 (33.6) |  |  |
| Yes | 76 (56.3) | 59 (43.7) |  |  |

**Table 6.** Associated Factors of Patient Safety Attitude on Stress Recognition.

| Domain | Stress Recognition |  | X<sup>2</sup> | p value |
| --- | --- | --- | --- | --- |
|  | Positive n(%) | Negative n(%) |  |  |
| Age (years) |  |  | 1.684 | 0.194 |
| Below 30 | 111 (53.1) | 98 (46.9) |  |  |
| 30 and above | 76 (46.3) | 88 (53.7) |  |  |
| Gender |  |  | 0.742 | 0.389 |
| Male | 30 (55.6) | 24 (44.4) |  |  |
| Female | 157 (49.2) | 162 (50.8) |  |  |
| Race |  |  | 2.130 | 0.144 |
| Malay | 139 (48.1) | 150 (51.9) |  |  |
| Non Malay | 48 (57.1) | 36 (42.9) |  |  |
| Education level |  |  | **12.331** | **0.002** |
| Diploma | 93 (43.5) | 121 (56.5) |  |  |
| Degree | 75 (56) | 59 (44) |  |  |
| Master/PHD | 19 (76) | 6 (24) |  |  |
| Position |  |  | **9.976** | **0.002** |
| Doctor | 86 (60.6) | 56 (39.4) |  |  |
| Nurse | 101 (43.7) | 130 (56.3) |  |  |
| Income |  |  | 0.132 | 0.716 |
| < RM 3860 | 93 (49.2) | 96 (50.8) |  |  |
| RM 3860 and above | 94 (51.1) | 90 (48.9) |  |  |
| Length of Working |  |  | 4.645 | 0.326 |
| Less than 1 year | 9 (39.1) | 14 (60.9) |  |  |
| 1-5 years | 98 (54.1) | 83 (45.9) |  |  |
| 6-10 years | 47 (52.2) | 43 (47.8) |  |  |
| 11-15 years | 18 (41.9) | 25 (58.1) |  |  |
| More than 15 years | 15 (41.7) | 21 (58.3) |  |  |
| Field of Working Area |  |  | **8.289** | **0.016** |
| Medical | 92 (56.8) | 70 (43.2) |  |  |
| Surgical | 55 (40.4) | 81 (59.6) |  |  |
| Others | 40 (53.3) | 35 (46.7) |  |  |
| History of Attending Safety Training |  |  | 0.031 | 0.860 |
| Yes | 118 (49.8) | 119 (50.2) |  |  |
| No | 69 (50.7) | 67 (49.3) |  |  |
| Number of Self- Reported Error |  |  | 1.854 | 0.173 |
| No (Zero Reporting) | 113 (47.5) | 125 (52.5) |  |  |
| Yes | 74 (54.8) | 61 (45.2) |  |  |

**Table 7.** Associated Factors of Patient Safety Attitude on Perception towards Management.

| Domain | Perception of Management |  |  |  |
| --- | --- | --- | --- | --- |
|  | Positive n(%) | Negative n(%) | X<sup>2</sup> | p value |
| Characteristic |  |  |  |  |
| Age (years) |  |  | **8.252** | **0.004** |
| Below 33 | 61 (29.2) | 148 (70.8) |  |  |
| 30 and above | 27 (16.5) | 137 (83.5) |  |  |
| Gender |  |  | 0.008 | 0.928 |
| Male | 13 (24.1) | 41 (75.9) |  |  |
| Female | 75 (23.5) | 244 (76.5) |  |  |
| Race |  |  | 2.885 | 0.089 |
| Malay | 74 (25.6) | 215 (74.4) |  |  |
| Non Malay | 14 (16.7) | 70 (83.3) |  |  |
| Education level |  |  | **16.604** | **0.000** |
| Diploma | 67 (31.3) | 147 (68.7) |  |  |
| Degree | 18 (13.4) | 116 (86.6) |  |  |
| Master/PHD | 3 (12) | 22 (88) |  |  |
| Position |  |  | **13.265** | **0.000** |
| Doctor | 19 (13.4) | 123 (86.6) |  |  |
| Nurse | 69 (29.9) | 162 (70.1) |  |  |
| Income |  |  | **10.700** | **0.001** |
| < RM 3860 | 58 (30.7) | 131(69.3) |  |  |
| RM 3860 and above | 30 (16.3) | 154 (83.7) |  |  |
| Length of Working |  |  | 3.768 | 0.438 |
| Less than 1 year | 9 (39.1) | 14 (60.9) |  |  |
| 1-5 years | 43 (23.8) | 138 (76.2) |  |  |
| 6-10 years | 18 (20) | 72 (80) |  |  |
| 11-15 years | 10 (23.3) | 33 (76.7) |  |  |
| More than 15 years | 8 (22.2) | 28 (77.8) |  |  |
| Field of Working Area |  |  | **19.131** | **0.000** |
| Medical | 24 (14.8) | 138 (85.2) |  |  |
| Surgical | 49(36) | 87 (64) |  |  |
| Others | 15(20) | 60 (80) |  |  |
| History of Attending Safety Training |  |  | **6.530** | **0.011** |
| Yes | 66 (27.8) | 171 (72.2) |  |  |
| No | 22( 16.2) | 114 (83.8) |  |  |
| Number of Self-Reported Error |  |  | 1.515 | 0.218 |
| No (Zero Reporting) | 61 (25.6) | 177 (74.4) |  |  |
| Yes | 27(20) | 108 (80) |  |  |

**Table 8.**
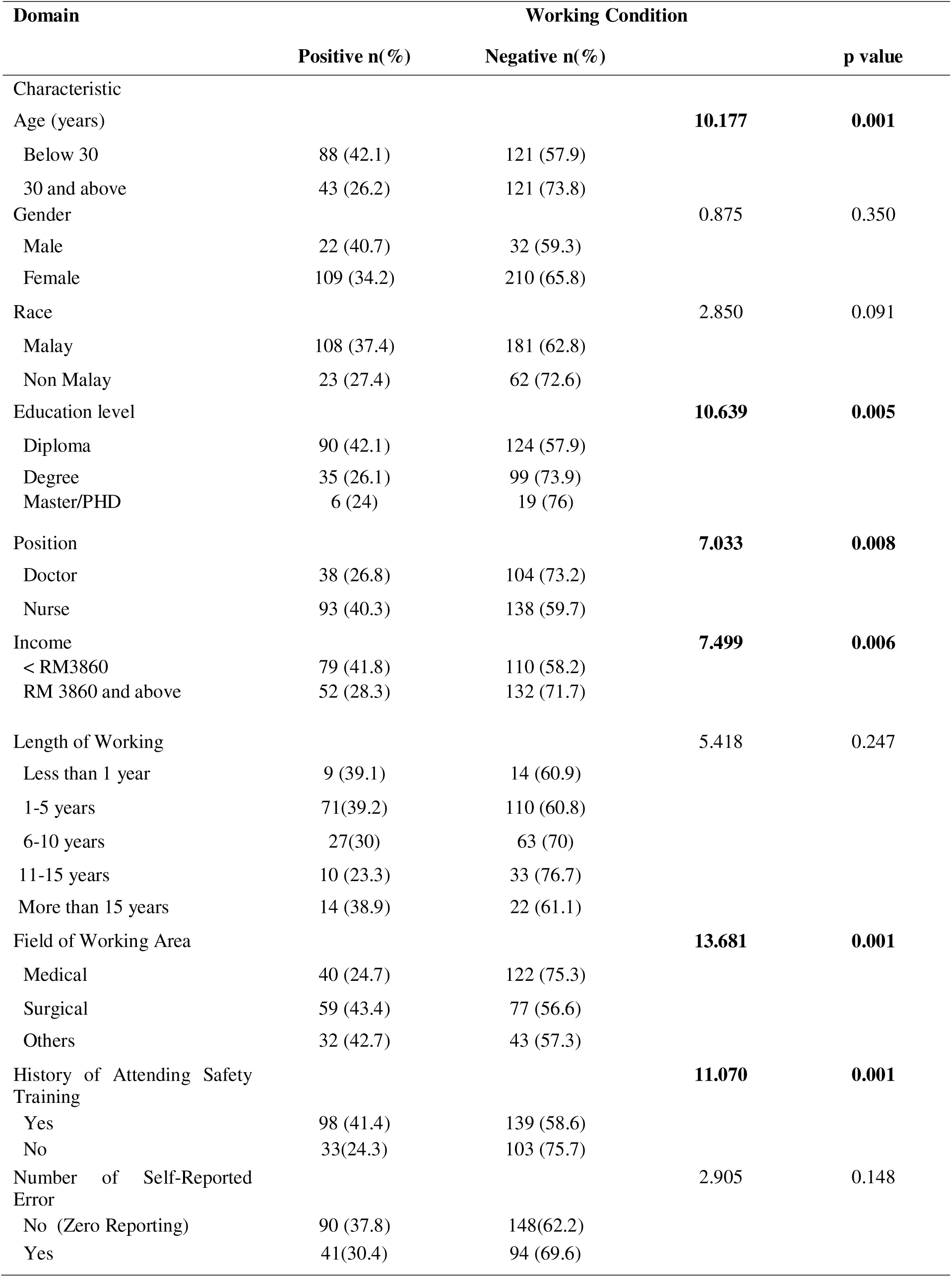
Associated Factors of Patient Safety Attitude on Working Condition.

**Table 9.** Logistic Regression Analysis for Factors Associated of Patient Safety Attitude Towards Teamwork.

| Domain | Teamwork |  |  |  |
| --- | --- | --- | --- | --- |
|  | Unadjusted OR<br>(95% CI) | P value | Adjusted OR<br>(95% CI) | P value |
| Characteristic |  |  |  |  |
| Position |  |  |  |  |
| Doctor | 1.661 (1.090-2.532) | **0.018** | 1.991(1.071-3.702) | **0.03** |
| Nurse | 1 |  |  |  |
| Income | 1 |  |  |  |
| < RM3860 |  |  |  |  |
| RM3860 and above | 1.553 (1.030-2.342) | **0.035** | 1.377 (0.771-2.459) | 0.279 |
| Field of Working Area |  |  |  |  |
| Medical | 0.470 (0.269-0.821) | **0.008** | 0.407(0.218-0.762) | **0.005** |
| Surgical | 0.467 (0.263-0.829) | **0.009** | 0.348(0.189-0.642) | **0.001** |
| Others | 1 |  |  |  |
| History of Attending Safety Training |  |  |  |  |
| Yes | 2.675(1.712-4.181) | **0.000** | 3.228 (1.973-5.282) | **0.000** |
| No | 1 |  |  |  |

For safety climate domain, the significant factors that has been identified in single logistic regression were field of working area and history of attending patient safety training. After running for multiple logistic regression, the determinants of having positive safety attitude towards safety climate were both work area and history of attending patient safety training (Table 4.10) From this analysis it can be interpreted as: 1) those who work in medical area have 2 times the odds to have negative attitude towards safety climate compared to non-clinical area (95% CI 0.257-0.892, *p*=0.020) and surgical area have 3 times the odds to have negative attitude towards safety climate compared to non-clinical area (95% CI 0.228-0.788, *p*=0.007) 2) those who attended patient safety training have 7 times the odds to have positive safety attitude towards safety climate compared to those who do not attend patient safety training (95% CI 4.076-13.012, *p*<0.0001).

**Table 10.** Logistic Regression Analysis for Factors Associated of Patient Safety Attitude Towards Safety Climate.

| Domain | Safety Climate |  |  |  |
| --- | --- | --- | --- | --- |
|  | Unadjusted OR<br>(95% CI) | P value | Adjusted OR<br>(95% CI) | P value |
| Characteristic |  |  |  |  |
| Field of Working Area |  |  |  |  |
| Medical | 0.349 (0.198-0.615) | **0.000** | 0.478(0.257-0.892) | **0.020** |
| Surgical | 0.530 (0.299-0.937) | **0.029** | 0.424(0.228-0.788) | **0.007** |
| Others | 1 |  |  |  |
| History of Attending<br>Safety Training |  |  |  |  |
| Yes | 7.194 (4.119-12.564) | **0.000** | 7.283 (4.076-13.012) | **0.000** |
| No | 1 |  |  |  |

**Table 11.** Logistic Regression Analysis for Factors Associated of Patient Safety Attitude Towards Job Satisfaction.

| Domain | Job Satisfaction |  |  |  |
| --- | --- | --- | --- | --- |
|  | Unadjusted OR<br>(95% CI) | P value | Adjusted OR<br>(95% CI) | P value |
| Characteristic |  |  |  |  |
| Race |  |  |  |  |
| Malay | 1.745(1.066-2.855) | **0.027** | 1.300 (0.729-2.317) | 0.373 |
| Non Malay | 1 |  |  |  |
| Education level |  |  |  |  |
| Diploma | 2.604 (1.122-6.043) | **0.026** | 35.709 (5.591-193.470) | **0.000** |
| Degree | 0.819 (0.348-1.926) | 0.647 | 1.209 (0.474-3.083) | 0.692 |
| Master/PHD | 1 |  |  |  |
| Position |  |  |  |  |
| Doctor | 0.460 (0.298-0.709) | **0.000** | 10.598 (3.043-36.905) | **0.000** |
| Nurse | 1 |  |  |  |
| Income |  |  |  |  |
| < RM3860 | 1.862 (1.217-2.849) | **0.004** | 0.533 (0.250-1.139) | 0.104 |
| RM3860 and above | 1 |  |  |  |
| Field of Working Area |  |  |  |  |
| Medical | 0.293 (0.159-0.541) | **0.000** | 0.383 (0.194-0.756) | **0.006** |
| Surgical | 0.845 (0.441-1.620) | 0.612 | 1.015 (0.495-2.084) | 0.967 |
| Others | 1 |  |  |  |
| History of Attending Safety Training |  |  |  |  |
| Yes | 3.158(2.030-4.913) | **0.000** | 2.883 (1.765-4.707) | **0.000** |
| No | 1 |  |  |  |

**Table 12.**
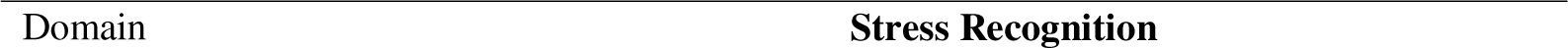
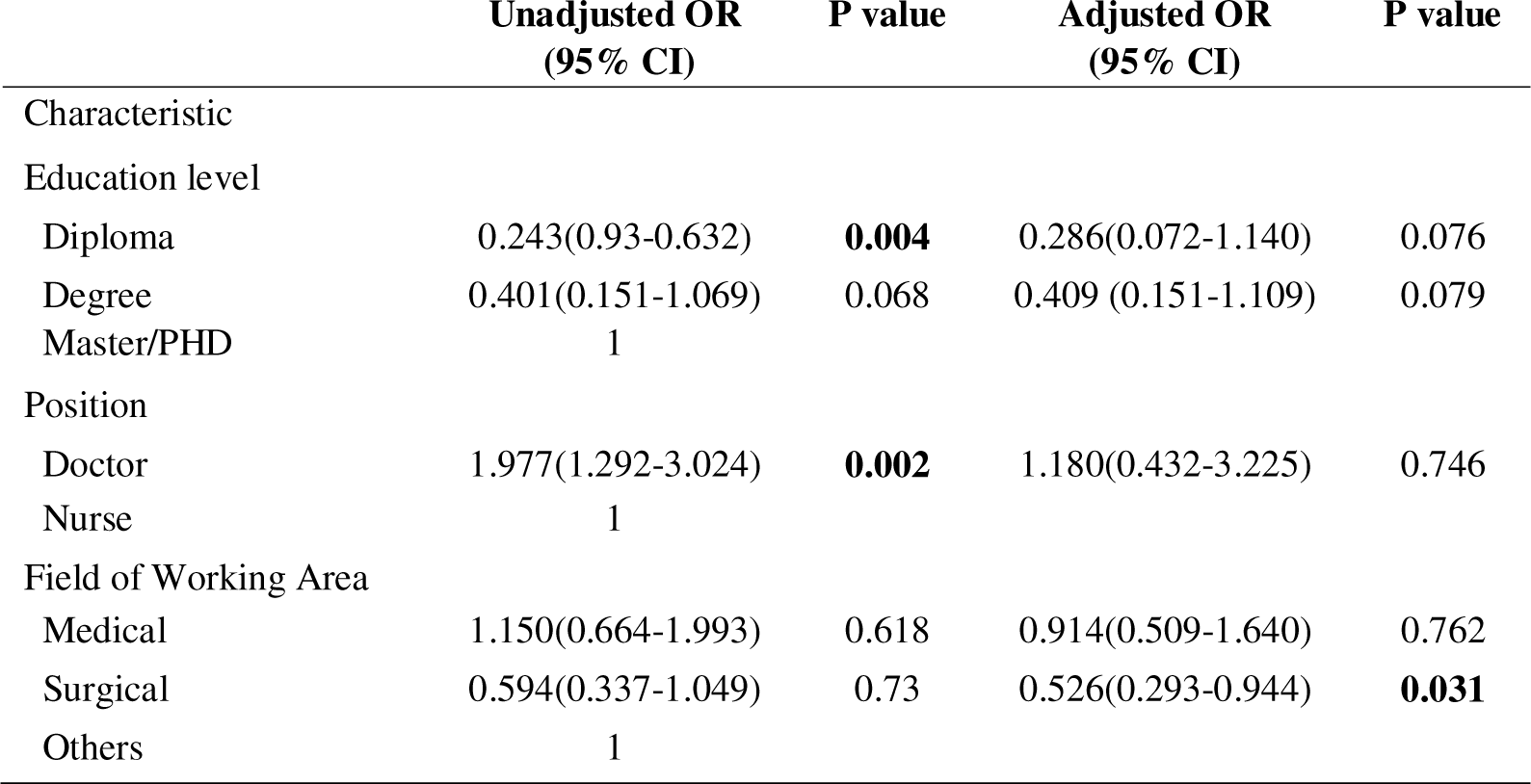
Logistic Regression Analysis for Factors Associated of Patient Safety Attitude Towards Stress Recognition.

| Domain | Stress Recognition |
| --- | --- |

| Characteristic | Unadjusted OR<br>(95% CI) | P value | Adjusted OR<br>(95% CI) | P value |
| --- | --- | --- | --- | --- |
| Education level |  |  |  |  |
| Diploma | 0.243(0.93-0.632) | **0.004** | 0.286(0.072-1.140) | 0.076 |
| Degree | 0.401(0.151-1.069) | 0.068 | 0.409 (0.151-1.109) | 0.079 |
| Master/PHD | 1 |  |  |  |
| Position |  |  |  |  |
| Doctor | 1.977(1.292-3.024) | **0.002** | 1.180(0.432-3.225) | 0.746 |
| Nurse | 1 |  |  |  |
| Field of Working Area |  |  |  |  |
| Medical | 1.150(0.664-1.993) | 0.618 | 0.914(0.509-1.640) | 0.762 |
| Surgical | 0.594(0.337-1.049) | 0.73 | 0.526(0.293-0.944) | **0.031** |
| Others | 1 |  |  |  |

**Table 13.** Logistic Regression Analysis for Factors Associated of Patient Safety Attitude Towards Perception of Management.

| Domain | Perception of Management |  |  |  |
| --- | --- | --- | --- | --- |
|  | Unadjusted OR<br>(95% CI) | P value | Adjusted OR<br>(95% CI) | P value |
| Characteristic |  |  |  |  |
| Age (years) |  |  |  |  |
| Below 30 | 2.091(1.257-3.480) | **0.005** | 2.084(1.120-3.878) | **0.021** |
| to be continued ... |  |  |  |  |
| ... continuation |  |  |  |  |
| 30 and above | 1 |  |  |  |
| Education level |  |  |  |  |
| Diploma | 3.342(0.967-11.555) | 0.057 | 4.064(0.564-29.313) | 0.164 |
| Degree | 1.138(0.309 - 4.194) | 0.846 | 0.754(0.185-3.0880) | 0.695 |
| Master/PHD | 1 |  |  |  |
| Position |  |  |  |  |
| Doctor | 0.363(0.207-0.634) | **0.000** | 1.642(0.412-6.541) | 0.482 |
| Nurse | 1 |  |  |  |
| Income |  | **0.001** | 0.805(0.352-1.842) | 0.608 |
| < RM3860 | 2.271(1.381-3.742) |  |  |  |
| RM3860 and above | 1 |  |  |  |
| Field of Working Area |  |  |  |  |
| Medical | 0.318(0.341-1.419) | 0.318 | 0.943(0.439-2.027) | 0.881 |
| Surgical | 2.253(1.158-4.382) | **0.017** | 2.461(1.222-4.960) | **0.012** |
| Others | 1 |  |  |  |
| History of Attending Safety Training |  |  |  |  |
| Yes | 2.000 (1.168-3.424) | **0.011** | 1.390(0.772-2.505) | 0.273 |
| No | 1 |  |  |  |

**Table 14.** Logistic Regression Analysis for Factors Associated of Patient Safety Attitude Towards Working Condition.

| Domain | Working Condition |  |  |  |
| --- | --- | --- | --- | --- |
|  | Unadjusted OR (95% CI) | P value | Adjusted OR (95% CI) | P value |
| Characteristic |  |  |  |  |
| Age (years) |  |  |  |  |
| Below 30 | 2.047 (1.314-3.188) | **0.002** | 2.200(1.297-3.732) | **0.003** |
| 30 and above | 1 |  |  |  |
| Education level |  |  |  |  |
| Diploma | 2.298 (0.883-5.985) | 0.880 | 3.857(0.737-20.190) | 0.110 |
| Degree | 1.120 (0.414-3.029) | 0.824 | 0.814(0.276-2.399) | 0.710 |
| Master/PHD | 1 |  |  |  |
| Position |  |  |  |  |
| Doctor | 1 |  |  |  |
| Nurse | 1.844 (1.170-2.908) | **0.008** | 0.403 (0.119-1.369) | 0.145 |
| Income |  |  |  |  |
| < RM3860 | 1.823(1.184-2.808) | **0.006** | 0.380 (0.380-1.600) | 0.780 |
| RM3860 and above | 1 |  |  |  |
| Field of Working Area |  |  |  |  |
| Medical | 0.441(0.247-0.787) | **0.006** | 0.509 (0.269-0.961) | **0.037** |
| Surgical | 1.030 (0.583-1.820) | 0.920 | 0.969 (0.528-1.780) | 0.919 |
| Others | 1 |  |  |  |
| History of Attending<br>Safety Training |  |  |  |  |
| Yes | 2.201 (1.376-3.520) | **0.001** | 1.738 (1.048-2.882) | **0.032** |
| No | 1 |  |  |  |

Race, education level, position in the hospital, income, field of working area and history of attending safety training were identified as the significant factors for job satisfaction domain in single logistic regression. But after running multiple logistic regression, the determinant factors of having positive safety attitude towards job satisfaction were only education level, position, work area and history of attending patient safety training. It can be interpreted as: 1) those workers with diploma education level have 36 times the odds to have positive safety attitude towards job satisfaction compared to master education level (95% CI 5.591-193.470, *p*<0.0001), 2) doctors have 11 times the odds to have positive attitude towards job satisfaction compared to nurses (95% CI 3.043-36.905, *p*<0.0001) 3) those who work in medical area have 3 times the odds to have negative attitude towards job satisfaction compared to non-clinical area (95% CI 0.194-0.756, *p*=0.006) 4) workers who attended patient safety training have 3 times the odds to have positive safety attitude towards job satisfaction compared to those who do not attend patient safety training (95% CI 1.765-4.707, *p*<0.0001)

For stress recognition, position and education level have been identified as the significant factors in single logistic regression. But after running multiple logistic regression, the determinant factor of having positive attitude towards stress recognition is only work area which; those who work in surgical area have 2 times the odds to have negative attitude towards stress recognition compared to non-clinical area (95% CI 0.293-0.944, *p*=0.031).

Age, position, income, field of working area and history of attending training were identified as the significant factors for perception of management domain in single logistic regression. After running multiple logistic regression, only age and work area were identified as the determinants factor of having positive safety attitude towards perception of management. From this analysis it can be interpreted as: 1) doctors and nurses with the ages of below 30 years old have 2 times the odds to have positive perception towards management compared to nurses (95%CI 1.120-3.878, *p*=0.021) 2) those workers who work in surgical area have 2 times the odds to have positive perception towards management compared to non-clinical area (95% CI 1.222-4.960, *p*=0.012).

The significant factors after running single logistic regression for safety attitude towards working condition were age, position, income, field of working area and history of attending patient safety training. But after went through multiple logistic regression, only age, work area and history of attending patient safety training were identified as the determinant factors of having positive attitude towards working condition. From this analysis, it can be interpreted as 1) workers with age below than 30 years old have 2 times the odds to have positive attitude towards working condition compared to workers with age more than 30 years old (95%CI 1.297-3.732, *p*=0.003) 2) doctors and nurses who works in medical area have 2 times the odds to have negative attitude towards working condition compared to nurses (95%CI 0.269-0.961,*p*=0.037) 3) those who have attended patient safety training have 2 times the odds to have positive attitude towards working condition compared to those who do not attend patient safety training (95% CI 1.048-2.882,p=0.32).

## DISCUSSION

In this study, we discussed on patient safety attitude among doctors and nurses who work at public hospitals in Kuala Lumpur and Selangor. Safety Attitude Questionnaire (SAQ) has been used in order to assess the patient safety attitude in this study. Patient safety is a discipline in the health care sector that applies safety science methods toward the goal of achieving a trustworthy system of health care delivery. Patient safety is also an attribute of health care systems; it minimizes the incidence and impact of, and maximizes recovery from, adverse events. [8]

Accumulating evidence supports the relationship between mature safety culture and patient safety, and improving a healthcare organization’s safety culture is associated with improved patient outcomes. The SAQ can be used to meet the increasing demand for safety culture assessment at the clinical area. Therefore, the Joint Commission on the Accreditation in the United Kingdom suggested hospitals should conduct safety culture surveys for safety improvement on the regular basis. These clinical areas benchmark their climate against other units in their institutions and against themselves.[6]

Many U.S hospitals have utilized valid questionnaires to measure safety attitudes among clinical areas and to compare changes in safety attitudes after evidence-based interventions. [9] Hospital safety managers can track the trends of culture changes of specific clinical units or the hospital. Strengths and weaknesses in the given clinical areas can be identified and appropriate interventions undertaken. For example, a poor teamwork climate would suggest collaborative rounds, whereas a poor safety climate would suggest Leadership Walk Rounds or a Comprehensive Unit-based Safety Program. [10,11] When used in a pre-intervention/post-intervention methodology, the SAQ factors have demonstrated sensitivity to quality improvement interventions at Kaiser Permanente [10] and recent evidence from Johns Hopkins Hospital demonstrates that climate can be targeted and improved.

The response rate of 83% achieved in this study is considered as a good response rate for studies on patient safety culture. The percentage is higher compared to the international benchmark of 66-72% [5], and other studies that used the same instrument, such as the study conducted in ICU in USA, 70.2% [12] another study in ambulatory setting in USA,69% [13] and yet one more among healthcare workers at several hospitals in Taiwan, 69.4% [14]. This might be due to the method of questionnaire administration used, whereby each respondent was given approximately 1-2 weeks time to return the survey, in order for them to really look into the questionnaire and answer them within given time; this suggest that the technique was effective in increasing response rates. On top of that, all managers in the involved hospitals supported the survey of patient safety culture. The SAQ itself has favourable characteristics; it is a self-reported anonymous questionnaire, the number of items is moderate, the wording is easy to understand, and the scoring method is simple and easy to learn. The survey required a short amount of time (10–15 min) to complete. A high response rate is an apparent sign of staff participation and attentiveness to quality issues, both signaling responsible behavior.

The attitude of safety addresses the matters of every person’s beliefs and his feelings about the safety issues and represents the sense of commitment and responsibility of the individual relation to safety issues. At the same time, it reflects a person’s belief in guidelines, policies, rules, procedures and safety procedures [15]. A desirable safety attitude in staff increases the safety level in the work environment, enhances the number of safe behaviors and reduces occupational accidents and near misses. An attitude of safety in each person indicates his previous readiness to provide a positive or negative reaction in response to the surrounding events, and at the same time, a combination of safe thinking and its tailored functioning .[16]

Since the attitude of a person forms the basis of his behavior, and, on the other hand, unsafe actions are known to be caused by factors such as negative and weak attitudes in the individual, measuring people’s attitude of safety can predict their safe or unsafe behaviors in the future. Moreover, through the use of appropriate controlling methods, the incidence of occupational accidents can be reduced and the safety of the work environment may be increased .[17] When comparing the mean score against national benchmark, our study scored higher for five out of the six safety domains: in decreasing score order, job satisfaction (73.8 vs 63.6), teamwork (70.7 vs 69.3), safety climate (69.6 vs 65.9), working condition(60.7 vs 57.2) and perception on management (59.0 vs 46.4) .[5]

Same goes with this study, some studies conducted in Palestine [18] Norway [19], USA[9] and Brazil [20] also found that job satisfaction scored the highest compared to other safety domains. There is also one study held in Malaysia involving cluster hospital also showed that job satisfaction scored the highest compared to other domains.[21] The positivity towards Job Satisfaction indicates that participants are reasonably satisfied with their job and will be positively involved towards accepting and implementing future Quality Improvement Initiatives. The importance of job satisfaction cannot be ignored because it is imperative, can improve staff enthusiasm, and stimulate productivity as well as quality of work. [18] Dissatisfaction may distract nurses and physicians from their patients, fail to provide proper care and increase turnover in the sector, which may subsequently jeopardize patients.

Perception of management received the lowest mean score from respondents. Management perception (attitude towards hospital management and ward support for the patient safety), which reflects the staff’s perception of management support in providing sufficient resources to create safe conditions. Developing a hospital safety culture involves complicated system engineering; this culture must be constructed based on initiation from upper management, and leadership roles must be strengthened to drive the general organization’s cognitions and behavior changes toward safety recognition. In recent years, continued attention has been given to patient safety in domestic and foreign medical industries Medical institutions in China are also committed to patient safety activities and have seen some advancements, while many patient safety projects focus on the improvement of unsafe factors in technology and procedures. [14,22]

Patient safety event reporting channels have been preliminarily established as part of the risk management system in hospitals in China since 2007, but the staff’s initiative and enthusiasm for reporting adverse events are affected by the punishment-based culture. [22] Additionally, an effective safety culture has not been attained because appropriate feedback regarding the staff’s job performance cannot be obtained, and open discussion of errors and accidents in the department cannot be facilitated. Promotion activities related to safety culture in organizations are not being performed or effectively internalized [41], and the medical staff’s attitudes, cognition, abilities, and behavior patterns regarding patient safety still need to be improved. Several studies reported the domain perception of management, received the lowest mean score as well .[ 5,20,23]

Stress recognition was scored below the international benchmark, demonstrating that respondents in this study had less positivity towards acknowledgement of how performance is influenced by stressors. It is also indicated that stress recognition is different from other dimensions of the SAQ scale because it requires the evaluation of one’s behavior in the workplace, while the other dimensions of the SAQ focus on others’ behavior. However, it is important to note that the stress recognition subscale does not contribute to the SAQ as intended an interpretation of results on this scale by its label ‘stress recognition’ may be misleading. [4,5] In our opinion, stress recognition can yield positive outcomes if respondents acknowledge the effects of stress on their performance and attribute it to desiring improved performance (eg, respondents with high stress recognition scores highlighted the need for increasing staffing levels); negative when they perceive it as an indicative of measuring their stress level and attributing it to suboptimal performance (eg, attributing it to increased frequencies of medication errors). Our opinion is also shared by Taylor and Pandian [24] who further suggested that this subscale be investigated for its true meaning.

One study done by Samsuri et al reported that teamwork climate was scored below international benchmark, demonstrating that respondents in their study had less positivity towards: input acceptance, speaking up, conflict resolution, feeling supported by others, ease of asking questions and collaboration with their own colleagues or other professionals. Working condition also reported scoring below international benchmark in their study demonstrating: negativity towards level of staffing, training of new personnel, availability of necessary information for therapeutic decision and supervision of trainees. [4]

The influence of teamwork should not be underestimated. The teamwork atmosphere, which deals with the interaction and communications of personnel, and represents the degree of trust, respect and mutual cooperation of individuals with each other. Many studies had linked a concrete relation between teamwork and patient safety with regards to communication and collaboration between unit team and teamwork climate. Accumulated findings have demonstrated the improvement in teamwork can significantly improve patient outcomes and decrease avoidable errors .[10] In the current complicated medical environment, healthcare providers have realized the importance of knowledge and complementary skills, resource sharing between team members, and establishing good cooperative relationships with colleagues to better manage conflict within the team with the clear target of ensuring patient safety. However, mutual trust between team members and the two-way communication capabilities in the department need to be improved, especially when perceiving a problem with patient care; speaking up is very important for patient safety, quality and efficiency in the patient treatment experience.

In our study, doctors and nurses with ages of below 30 years old have more positive attitudes toward working condition and perception of management. In contrast, Oi Ling Siu et al. found that some older workers do have more positive attitudes towards safety, compared with younger workers .[ 25] Based on study done in Norway, older respondents and those who spend more time at work might feel more comfortable in the nursing home clinical setting. On the other hand, younger employees may be a group that identifies possible risks easier. They are more recently trained and have probably been introduced to the issue of patient safety to a wider degree than their older colleagues. This may have created increased patient safety awareness, which is reflected in the lower scores in this group and may explain why several patient safety factors are perceived as being poorer among the younger employees .[ 26]

In line with Rigobello et al, we also found that there is no significant difference for all domains in patient safety attitude between male and female workers. However, in a study conducted in the US on 187 nurses working in the operating room, collated results meaningfully suggested a higher positive safety attitude in women than the men .[ 27] One of the reasons for such difference in outcome can be attributed to the contribution of male and female nurses. However, women are generally more sensitive to keeping with the safety, quality of care towards patient as well as the use of safety principles .[28]. According to another study in Rio also said female professionals prevail in their study due to the construction of gender roles, which direct the professional choice influenced by family, social history, conditioning childhood ideas, behavioural stereotypes and a career choice consistent career with femininity .[29]

According to our study, doctors have more positive attitude towards teamwork and job satisfaction. Our findings appear to be in line with previous study, Indre et al. found physicians had significantly higher safety attitudes related to teamwork climate, stress recognition and perception of management than nurses in the group of health care professionals who also did not report a safety incident. However, our results indicate there is no association between self-reported incidents and patient safety attitude. [13]. Modak et al said there were some notable differences in scores among types of providers. Very few physicians had positive assessments of management, in stark contrast to the percentage of managers having a positive perception of themselves. The other statistically significant finding was that nurses had the highest stress recognition scores and medical assistants the lowest. Higher stress recognition scores indicate more recognition of the effects of stress on the ability of a provider to perform optimally in delivering safe care. Although not statistically significant, managers and nurses had the highest safety attitude scores .[13] This is consistent with other research that consistently reports better attitudes from those at the top of the hierarchy in organizations. While it can be helpful for leaders to have positive attitudes, it may be a problem if their attitudes reflect an unrealistic view of the practice or if their attitudes are markedly different from those of others .[11]

In a study undertaken at obstetric centres in the USA, statistically significant associations were found between the domain teamwork climate and professional category, with physicians showing better scores than nurses.. That study demonstrated different viewpoints and opinions among physicians and nurses regarding safety issues, with physicians demonstrating greater awareness in reports on potential damage when compared to nurses. [31]

Tirgar et al. found a significant relationship was between education and safety attitude. The chance for nurses with education levels higher than a bachelor’s degree in gaining safety attitude was 30.3 times higher than that of nurses with a bachelor’s degree .[27] In contrast, our study found those workers with diploma education seemed to have more positive attitude in job satisfaction compared to other education level. Such difference in outcome can be attributed to the proportion between nurses and doctors who participated in this study. Nurses who contributed about 60% of all respondents mainly have diploma education level. Moreover, job satisfaction domain scored the highest percentage among nurses compared to other domains.

Several studies have relied that length of working experience as major indicator for their safety attitude. Bianca et al in his study has established that professionals with 21 years of experience or more presented better safety attitude when compared to the other professionals. [20] But anyway, in this study, there is no association between length of working experience and the employees’ attitude towards patient’s safety. A significant relationship was found between work experience and the employees’ attitude towards patient’s safety. Thus, the safety attitude of nurses with work experience between 10 and 20 year and nurses with a history of over 20 year were respectively estimated 2 and 4 times more than the less experienced nurses (less than 10 year) .[27] Other several studies in nurses also suggest the issue, which mainly relates to gaining more work experience and the formation of a positive safety culture and attitude during the years of working. [32]

Except for stress recognition, those who work in medical area tend to have negative safety attitude towards all safety domains compared to other working area. Prior study also shown for the variable professional activity (whether care and non-care), the associations were significant for the domain perception of unit and hospital management (p=0.01), where the non-care professionals obtained a better score than the professionals active in direct care. Authors suggest that the better perception among management than among care professionals can be explained by the managers’ sense of propriety and responsibility regarding their roles in the hospital infirmaries .[20] Study done by Samsuri et al. revealed that, for these domains teamwork climate, safety climate and job satisfaction, and overall safety culture, pharmacists working in outpatient and ambulatory care reported significantly higher scores than those working elsewhere. This could be explained by the proportion of pharmacists in outpatient and ambulatory unit, which is normally higher than that in other units such as inpatient and clinical. Therefore, those pharmacists have more opportunities for interacting with their peers within the same unit, while being minimally involved in collaborative activities with other healthcare professionals. As a result, they have better attitudes towards teamwork, safety and job satisfaction. [4]

Meanwhile, the multidisciplinary nature of the job in inpatient and in clinical settings would mean that the pharmacists would need to interact and build a good rapport with other healthcare professionals. Job conflicts may occur daily, which may influence pharmacists’ satisfaction and positive perception on teamwork and safety climate .[33] Sexton et al also recognised the scores of teamwork climate to be higher within a group of peers.[5]

In this study, history of attending patient safety training seems to have the most significant relationship with patient safety attitude domains except for stress recognition and perception of management. Previous study also suggested that changes in work-related attitudes are important outcomes of training. Many researchers show that employee attitude toward patient safety is positively associated with participation in patient safety training courses. The more trainings attended, the more knowledge about safety will direct their perception towards safety attitude. Regarding more specific training, Cabrera et al found that participation in safety training produces a positive attitude toward safety and a significant relationship between employees’ safe behavior and both the climate of safety and attitudes toward safety .[ 34]

By other study, attitudes have been found to be more positive after training, and similar to the improvements of knowledge reported by Brasaite et.al [30] The same study showed that after a day training course on patient safety, senior doctors’ safety attitudes had significantly improved post course and were sustained based on their own evaluations. In the western part of Lithuania, nurses did not pass the patient safety training had more negative safety attitude, which covers the present study results. [30] Therefore, doing measures such as training need assessment and using it to hold personnel and patient safety courses and workshops according to new scientific and research findings is very useful and effective in increasing the nurses’ safety attitude as well as in enhancing the satisfaction and improving the quality of patient care and preventing the occurrence of incidents in this profession group.

Study done by Indre et al, who found that in the health care professional group who had reported a safety incident during last year, had significantly higher safety attitudes related to their teamwork climate. Those who did not report any safety incidents during the last year had more positive attitudes towards stress recognition than those who had reported such incidents [30]. But in this study, there is no association between number of self-reported events and the employees’ attitude towards patient’s safety.

Previous studies had also found that many errors in healthcare were under-reported due to possible barriers such as having busy working schedules, severity of patient harm and anxieties about harming interprofessional .[35] Other correlation studies from the USA also concluded that safety culture influenced the occurrence of medication errors and adverse events, where, in a positive environment, staff were less likely to commit an error and an adverse event was less likely to occur .[36]

## CONCLUSION

This study showed doctors and nurses at public hospitals in Kuala Lumpur and Selangor scored higher mean score in almost all patient safety domains compared to international benchmark. Each domain seemed to have different determinant factors except for history of attending safety training. There is a significant association between this factor with all patient safety attitude domains except for stress recognition and perception of management.

The identification of the predictors of patient safety scores is an important tool that, linked with organizational actions, permits diagnosing, intervening and executing activities, based on the domains that need to be improved (work conditions and management actions) and the professionals’ intrinsic and extrinsic factors in need of attention (stress, teamwork and satisfaction). All these efforts contribute to implementation of safety climate at the institution, with the promotion of patient safety as the final result.

Overall result of this study illustrated that our study scored higher for five out of the six safety domains when comparing the mean score against national benchmark except for stress recognition. Stress recognition was scored below the international benchmark, demonstrating that respondents in this study had less positivity towards acknowledgement of how performance is influenced by stressors. It is also indicated that stress recognition is different from other dimensions of the SAQ scale because it requires the evaluation of one’s behavior in the workplace, while the other dimensions of the SAQ focus on others’ behavior.

From this study we found that the determinant factors of having positive safety attitude towards job satisfaction were only education level, position, work area and history of attending patient safety training. While position, work area and history of attending patient safety training were the determinants factors for having positive safety attitude towards teamwork. Only work area and history of attending patient safety training were the determinants of having positive safety attitude towards safety climate. For stress recognition, the determinants factor of having positive attitude towards stress recognition is only work area. Age and work area were identified as the determinants factor of having positive safety attitude towards perception of management. And lastly, the determinant factors of having positive attitude towards working condition were age, work area and history of attending patient safety training.

Result from this study also showed that each domain of patient safety seemed to have different determinant factors except for history of attending safety training. There is a significant association between this factor with all patient safety attitude domains except for stress recognition and perception of management. Doctors and nurses with history of attending patient safety attitude have more positive attitude towards patient safety. However, this factor is not significant for stress recognition and perception of management.

## RECOMMENDATION

- The safety culture in most hospitals is not fully established and needs to be improved through intervention strategies. To insert the concept of “first, do no harm” into each unit and every operation code in the healthcare organization, we should accurately understand and grasp the connotation of a safety culture and its basic elements, highlight the important influence of human factors on the safety of patients, emphasize the important role of safety culture at every management level, and constantly improve communication and cooperation within the team and between teams.
- Promotion activities related to safety culture in organizations should be performed or effectively internalized and the medical staff’s attitudes, cognition, abilities, and behaviour patterns regarding patient safety still need to be improved.
- Patient safety training (Workshop, Courses, Seminar and CME) should be continued and regularly done to all healthcare workers to make sure every employee has been exposed to patient safety especially to those who directly involved with patient care.
- Executive Walk Rounds (EWRs) should be introduced in public hospitals because EWRs enlist leadership to breakdown the significant barriers to discussion of human error in healthcare and help hospitals identify opportunities to improve care processes. These rounds demonstrate the executives and the organization’s commitment to patient safety, and may improve provider attitudes regarding safety-related issues.
- Policymakers need to ensure that legislations and regulations are introduced to encourage healthcare organizations to implement patient safety reporting systems which will help identify risks to patients and help healthcare organizations learn from their mistakes.
- Safety culture should be assessed on a regular basis to evaluate the effectiveness of patient safety programs and interventions.

## LIMITATIONS

This study was one of the few studies in an effort to evaluate the patient safety attitude among healthcare workers. Most of other study worldwide used bigger sample to evaluate the patient safety attitude. The factor of limited time and financial constraint limit the ability to include bigger sample into this study. Another drawback is that we did not explore the connection between patient safety attitude and the patient outcome. Further research is required to identify the complicated relationship between patient safety culture and incident reporting system, the number of reporting, patient outcome and how the data produced can be translated into action and learning points. The findings are crucial and can guide us in interventions and improvements to create a safe healthcare system and to reduce adverse medical outcomes. The use of a questionnaire to evaluate safety attitude or a particular safety environment plays an essential role in planning the evaluation of an institution’s safety culture. Although a useful tool, SAQ has its limitation; it assesses staffs’ beliefs regarding the safety culture, rather than their real safety behaviour .[37] Notably, SAQ tests the current attitude regarding patient safety; however, there may be differences between attitudes and actual practice. Therefore, to explore the dimensions that influence patient safety in more detail, SAQ should be combined with qualitative methods such as peer observation, group discussions, analysis of organisation’s incident history and audits of the safety management system. [38,39,40]. A wide gap in research remains to how data obtained from different methods are related and how to combine them to get a complete safety culture view. Despite these limitations, we believe this research offers useful insight into our organisation’s baseline patient safety attitude.

## Data Availability

All data produced in the present study are available upon reasonable request to the authors

