## APPENDIX 2 for "ASSESSMENT OF PATIENT SAFETY ATTITUDE AMONG DOCTORS AND NURSES AT PUBLIC SPECIALIST HOSPITALS IN MALAYSIA"

**APPENDIX D**

### <u>INFORMED CONSENT FORM</u>

**RESEARCH TITLE:** Assessment On Patient Safety Attitude Among Doctors And Nurses At Public Hospitals In Kuala Lumpur And Selangor

**Researcher's Name: Dr Sharifah Balqis binti Sayed Abdul Hamid**

I, ……………………………………..............IC No : ………….............................

- have read the information in the Respondent Information Sheet **including information regarding the risk in this study.**
- have been given time to think about it and all of my questions have been answered to my satisfaction.
- understand that I may freely choose to withdraw from this study at anytime without reason and without repercussion
- understand that my anonymity will be ensured in the write-up.

I voluntarily agree to be part of this research study, to follow the study procedures, and to provide necessary information to the researcher, as requested.

………………………………. ……………………..

(Signature) (Date)

| | |
|---|---|
| ………………………………. | ………………………………. |
| Witness (if any) | Researcher |
| ………………………………. | ………………………………. |
| (Signature) | (Signature) |
| ………………………………. | ………………………………. |
| (IC Number) | (IC Number) |
| ………………………….…. | ………………………………. |
| (Date) | (Date) |

**APPENDIX E**

#### INFORMATION SHEET

**Research Title:** Assessment On Patient Safety Attitude Among Doctors And Nurses At Public Hospitals In Kuala Lumpur And Selangor

**Introduction:**

You are most welcome to participate in this research study. Before that, it is important that you take your time to read and understand the information of the research in this Information Sheet.

**Purpose of Study:**

Ministry of Health has started implementing Malaysian Patient Safety Goals in healthcare facilities since 2013 to improve our Patient Safety Culture. Good Patient Safety Culture indicated by the better the perception of Patient Safety Attitude among the healthcare providers. This study aims to access Patient Safety Attitude among doctors and nurses who works in public hospitals in Kuala Lumpur and Selangor.

**What will the study involve?**

The study will require you to answer questionnaire regarding Patient Safety Attitude according to six domains which is teamwork, safety climate, job satisfaction, stress recognition, perception toward management and working condition that make up the patient safety attitude. This questionnaire will also be asking your demographic details, number of self-reported events and history of patient safety training attended.

**Risks and Benefits:**

By participating in this research, you are required to state your perception on various aspect that can determine successfulness of the adoption. This indirectly give you a channel to voice your view to the stakeholder. Therefore, remedial action can be taken in order to improve Patient Safety Culture .

There is no risk either directly or indirectly that may occur if you are participated in this research.

**How Many People Will Participate?**

Approximately 400 participants will take part in this study.

**How Long Will I Be In This Study?**

This will take no longer than 20 minutes to complete

**Do you have to take part?**

Participation in this study is totally voluntary. Once you agree to participate, then you will be asked to sign the “Informed Consent Form”. The copy of "Informed Consent Form" and this Information Sheet will be given to you for your reference.

Once you have decided to participate, you can still withdraw from the study without penalty.

Your data will not be used and will be discarded. The same concept applied that the researcher may also remove you from the study for a variety of reason.

**Data & Confidentiality:**

The data derived from this research will be transformed into a report which may be published. Access to the data is only by the research team. The data will be reported in a collective manner with no reference neither directly nor indirectly to any particular individual. Hence your identity will be kept confidential.

**Payment and compensation:**

You do not have to pay nor will you be paid to participate in this study.

**Who can I ask about the study?**

If you have any queries, you can direct them to the research officer. You can also contact the UKM Department of Community Health for clarifications.

Research Officer

Dr Sharifah Balqis binti Sayed Abdul Hamid

Department of Community Health

UKM Medical Centre

Phone Number : 03-9145 5887 / 5888 / 5889

Mobile : 013-9325242
