## APPENDIX A for "ASSESSMENT OF PATIENT SAFETY ATTITUDE AMONG DOCTORS AND NURSES AT PUBLIC SPECIALIST HOSPITALS IN MALAYSIA"

### SAFETY ATTITUDE QUESTIONAIRE

#### SECTION A : BACKGROUND INFORMATION

Please select ONLY one answer

Age (Year) : ☐<30 ☐31-40 ☐ 41-50 ☐ >50

Gender : ☐Female ☐Male

Race : ☐Malay ☐ Chinese ☐ Indian ☐ Others

Education Level : ☐Diploma

☐ Degree

☐ Master&PHD

Salary : ☐<RM3860 ☐ RM3860 -RM8319 ☐>RM8319

SECTION B : WORK BACKGROUND INFORMATION

Position : ☐Doctor ☐ Nurses

Length of Working Experiences (years) : ☐ < 1

☐ 1-5

☐ 6-10

☐ 11-15

☐ >15

Field of Working Area : ☐ Medical

☐ Surgical

☐ Others

### SECTION D : FRONTLINE PERSPECTIVES FROM THIS PATIENT CARE AREA (SAQ)

Please answer the following items with respect to your specific unit or clinical area. Choose your responses using the scale below:

☐1 Disagree strongly
☐2 Disagree slightly
☐3 Neutral / Neutral
☐4 Agree slightly
☐5 Agree Strongly

1. Nurse input is well received in this clinical area. ☐1 ☐2 ☐3 ☐4 ☐5

2. In this clinical area, it is difficult to speak up if I perceive a problem with patient care. ☐1 ☐2 ☐3 ☐4 ☐5

3. Disagreements in this clinical area are resolved appropriately (i.e., not *who* is right, but *what* is best for the patient). ☐1 ☐2 ☐3 ☐4 ☐5

4. I have the support I need from other personnel to care for patients ☐1 ☐2 ☐3 ☐4 ☐5

5. It is easy for personnel here to ask questions when there is something that they do not understand ☐1 ☐2 ☐3 ☐4 ☐5

6. The physicians and nurses here work together as a well-coordinated team. ☐1 ☐2 ☐3 ☐4 ☐5

7. I would feel safe being treated here as a patient. ☐1 ☐2 ☐3 ☐4 ☐5

8.Medical errors are handled appropriately in this clinical area. ☐1 ☐2 ☐3 ☐4 ☐5

9.I know the proper channels to direct questions regarding patient safety in this clinical area. ☐1 ☐2 ☐3 ☐4 ☐5

10. I receive appropriate feedback about my performance. ☐1 ☐2 ☐3 ☐4 ☐5

11. In this clinical area, it is difficult to discuss errors. ☐1 ☐2 ☐3 ☐4 ☐5

12. I am encouraged by my colleagues to report any patient safety concerns I may have. ☐1 ☐2 ☐3 ☐4 ☐5

13. The culture in this clinical area makes it easy to learn from the errors of others. ☐1 ☐2 ☐3 ☐4 ☐5

14. My suggestions about safety would be acted upon if I expressed them to Management. ☐1 ☐2 ☐3 ☐4 ☐5

15. I like my job. ☐1 ☐2 ☐3 ☐4 ☐5
16. Working here is like being part of a large family. ☐1 ☐2 ☐3 ☐4 ☐5
17. This is a good place to work. ☐1 ☐2 ☐3 ☐4 ☐5
18. I am proud to work in this clinical area. ☐1 ☐2 ☐3 ☐4 ☐5
19. Morale in this clinical area is high. ☐1 ☐2 ☐3 ☐4 ☐5
20. When my workload becomes excessive, my performance is impaired. ☐1 ☐2 ☐3 ☐4 ☐5
21. I am less effective at work when fatigued. ☐1 ☐2 ☐3 ☐4 ☐5
22. I am more likely to make errors in tense or hostile situations. ☐1 ☐2 ☐3 ☐4 ☐5
23. Fatigue impairs my performance during emergency situations (e.g. emergency resuscitation, seizure). ☐1 ☐2 ☐3 ☐4 ☐5
24. Management supports my daily efforts. ☐1 ☐2 ☐3 ☐4 ☐5
25. Management doesn't knowingly compromise patient safety. ☐1 ☐2 ☐3 ☐4 ☐5
26. Management is doing a good job. ☐1 ☐2 ☐3 ☐4 ☐5
27. Problem personnel are dealt with constructively by our management. ☐1 ☐2 ☐3 ☐4 ☐5
28. I get adequate, timely info about events that might affect my work, from management. ☐1 ☐2 ☐3 ☐4 ☐5
29. The levels of staffing in this clinical area are sufficient to handle the number of patients. ☐1 ☐2 ☐3 ☐4 ☐5
30. This hospital does a good job of training new personnel. ☐1 ☐2 ☐3 ☐4 ☐5
31. All the necessary information for diagnostic and therapeutic decisions is routinely available to me. ☐1 ☐2 ☐3 ☐4 ☐5
32. Trainees in my discipline are adequately supervised. ☐1 ☐2 ☐3 ☐4 ☐5
33. I experience good collaboration with nurses in this clinical area. ☐1 ☐2 ☐3 ☐4 ☐5
34. I experience good collaboration with staff physicians in this clinical area. ☐1 ☐2 ☐3 ☐4 ☐5
35. I experience good collaboration with pharmacists in this clinical area. ☐1 ☐2 ☐3 ☐4 ☐5
36. Communication breakdowns that lead to delays in delivery of care are common. ☐1 ☐2 ☐3 ☐4 ☐5

**SECTION E: NUMBER OF SELF-REPORTED EVENTS**

In the past 12 months, how many event reports have you filled out and submited?

An “event” is defined as any type of eror, mistake, incident, accident, or deviation, regardless of whether or not it results in patient harm.

☐ No events reported
☐ 1-10 events reported
☐ >10 events reported

**SECTION E : HISTORY OF PATIENT SAFETY TRAINING ATTENDED**

Have you attend any Patient Safety Training during your work experience?

☐Yes ☐ No

**Thank you for completing this survey - your time and participation are greatly appreciated.**
